## Supplement for "Importation of Alpha and Delta variants during the SARS-CoV-2 epidemic in Switzerland: phylogenetic analysis and intervention scenarios": suplement_voc_imports.pdf

**Supporting information**

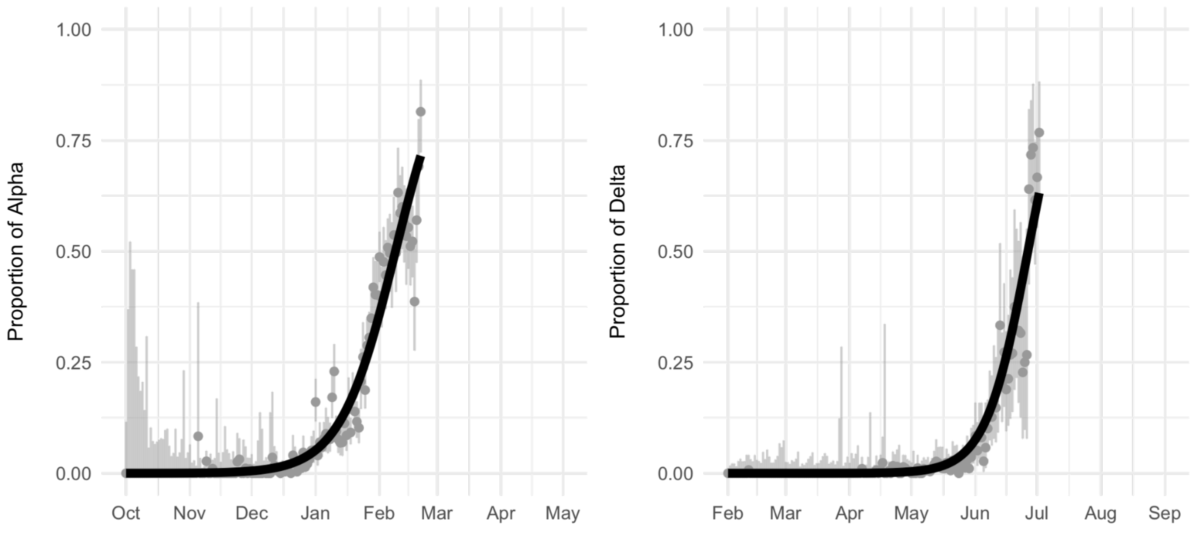


**Sup Fig 1.** Fit of the logistic growth model to the proportion of SARS-CoV-2 VoCs. Gray dots: Genomic surveillance data with 95% binomial confidence interval. Black line: Model fit.

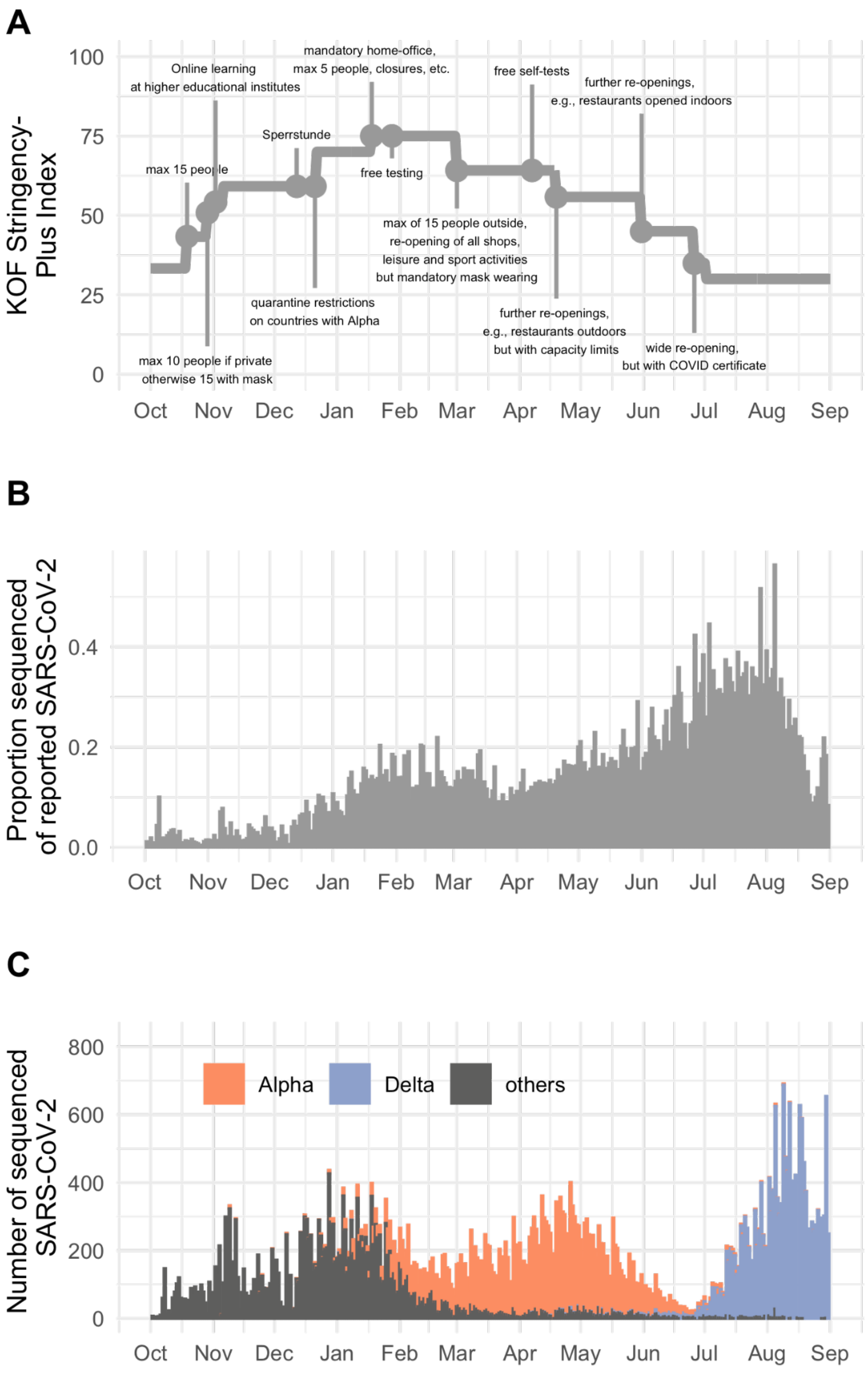


**Sup Fig 2:** Measures and genomic sequencing during the SARS-CoV-2 epidemic in Switzerland from October 2020 to September 2021. A: The KOF stringency plus index recorded the stringency of SARS-CoV-2 policy measured in Switzerland over time. The values range from 0 (= no measures) to 100 (= full lockdown). B: Proportion of reported SARS-CoV-2 cases that were sequenced. **C**: Number of sequenced SARS-CoV-2 cases.

**Sup Table 1.** Estimated importation of VoCs and simulated impact on the SARS-CoV-2 epidemic in Switzerland. Abbreviation: VoC, variant of concern.

| | **Alpha** | | **Delta** | |
|---|---|---|---|---|
| | **Liberal** | **Conservative** | **Liberal** | **Conservative** |
| **Estimated imports from the phylogeny** | 1,038 | 383 | 1,347 | 455 |
| **Simulation period** | 1 Oct 2020 - 1 May 2021 | | 1 Feb 2021 - 1 Sept 2021 | |
| **Total simulated reported cases** | 593,418 | 592,768 | 288,397 | 271,702 |
| **Number of simulated variant cases** | 97,116 (16%) | 70,898 (12%) | 110,596 (38%) | 87,861 (32%) |
| **Date by which 50% of cases are the variant (dominance)** | 05 Mar 2021 | 22 Mar 2021 | 30 Jun 2021 | 09 Jul 2021 |

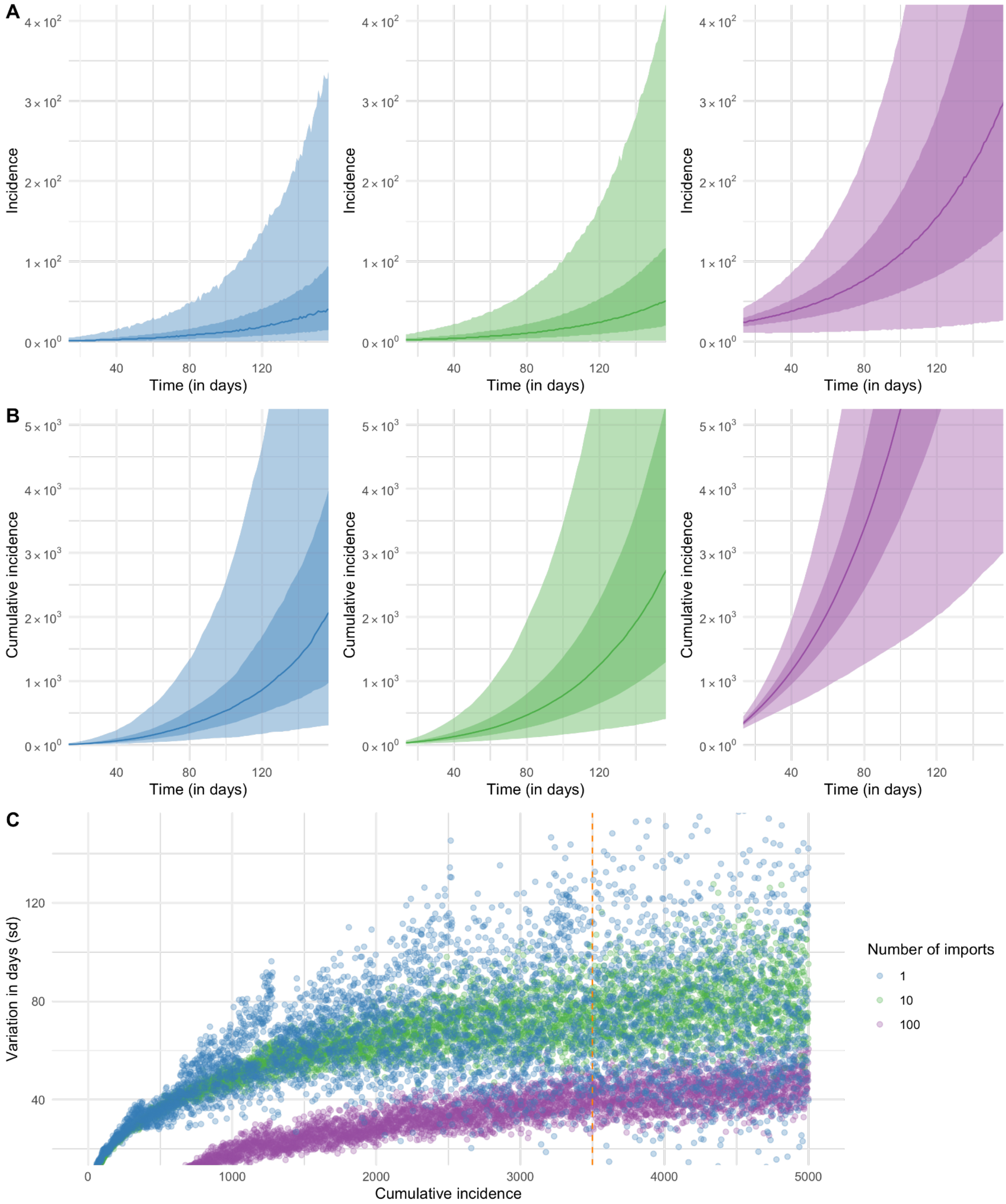


**Sup Fig 3:** Stochastic effects during the early growth phase of SARS-CoV-2 variants. A, B: Time to reach a certain (cumulative) incidence. Shaded regions correspond to the 50% and 95% interval of all simulations. C: Variation in the time to reach a certain cumulative incidence expressed as standard deviation.
